# Prognostic Value of Post-Procedural Stress Hyperglycemia Ratio in Patients Undergoing Percutaneous Coronary Intervention: Insights Across Glucose Metabolic Status

**DOI:** 10.1101/2025.10.28.25338989

**Authors:** Queyun Sun, Cheng Cui, Lin Jiang, Jingjing Xu, Yi Yao, Na Xu, Xiaozeng Wang, Zhenyu Liu, Weiting Cai, Zheng Zhang, Yongzhen Zhang, Xiaogang Guo, Zhifang Wang, Yingqing Feng, Qingsheng Wang, Jianxin Li, Xueyan Zhao, Jue Chen, Runlin Gao, Lei Song, Yaling Han, Jinqing Yuan, Ying Song

## Abstract

**Background:** The stress hyperglycemia ratio (SHR) is a novel marker reflecting true acute hyperglycemic status and has been linked to adverse clinical outcomes in various settings. However, the clinical significance of SHR measured after percutaneous coronary intervention (PCI) remains inadequately explored.

**Objective:** To investigate the relationship between post-procedural SHR and long-term prognosis in patients with coronary artery disease (CAD).

**Methods and Results:** In this prospective cohort study, a total of 7,423 patients with complete baseline data were included. Kaplan-Meier survival analysis showed significant differences in the incidence of all adverse outcomes across SHR tertiles during a 2-year follow-up (Log-rank P < 0.05). In multivariable Cox regression, patients in the highest SHR tertile had an independently increased risk of major adverse cardiovascular events (MACE) [adjusted HR (95% CI): 1.254 (1.046–1.502), P = 0.014], mainly attributable to higher risks of all-cause death [1.531 (1.076–2.177), P = 0.018] and cardiac death [1.702 (1.111–2.607), P = 0.014]. Restricted cubic spline analysis revealed a J-shaped association between SHR and the risk of MACE. Stratified analysis found that the effect of post-procedural SHR on prognosis was observed in patients with diabetes, regardless of the guideline-based criteria used to define glucose metabolism status, whereas no significant association was observed in those with normoglycemia or prediabetes.

**Conclusion:** Elevated post-procedural SHR is independently associated with a higher risk of adverse cardiovascular outcomes in patients with CAD, particularly among those with diabetes. Assessment of post-procedural SHR may help refine risk stratification and guide individualized management in this population.

## 1. Introduction

Stress hyperglycemia refers to a transient elevation of blood glucose levels during acute stress conditions, reflecting the neuroendocrine response and the degree of insulin resistance under stress [1].

Traditional glycemic indicators, such as random or admission glucose, are easily influenced by baseline glucose metabolism and therefore cannot accurately reflect the magnitude of the stress response. The stress hyperglycemia ratio (SHR), defined as the ratio of admission glucose to the estimated average glucose derived from glycated hemoglobin (HbA1c), effectively quantifies the acute rise in glucose relative to an individual’s chronic glycemic background [2].

Accumulating evidence has demonstrated that an elevated stress hyperglycemia ratio (SHR) is strongly associated with adverse clinical outcomes across a wide range of conditions, including atherosclerotic cardiovascular diseases [3-6], heart failure [7, 8], chronic kidney disease [9], stroke [10, 11], or critical illness [12-14].

However, most existing studies have primarily focused on the prognostic implications of admission SHR values, whereas the prognostic value of post-procedural SHR in patients undergoing percutaneous coronary intervention (PCI) remains underexplored. Although PCI significantly improves coronary perfusion and clinical outcomes, it is an important physiological stressor characterized by inflammatory cascades [15], reperfusion injury [16], and platelet activation [17, 18]. Persistent hyperglycemia following revascularization may reflect ongoing metabolic imbalance and insulin resistance, which could have independent prognostic implications.

Moreover, whether the prognostic impact of post-procedural SHR differs among patients with varying glucose metabolism status—namely normoglycemia, prediabetes, and diabetes—remains unclear.

Therefore, the present study aims to systematically evaluate the association between post-procedural stress hyperglycemia ratio (SHR) and the risk of major adverse cardiovascular events (MACE) in patients undergoing PCI, and to explore the potential heterogeneity of this association across different glucose metabolism strata.

## 2. Methods

### 2.1. Study cohort

This study utilized data from the PRospective Observational Multicenter cohort for Ischemic and hEmorrhage risk in coronary artery disease patients (the PROMISE cohort), which enrolled 18,701 hospitalized patients across nine centers in China between January 2015 and May 2019. Inclusion criteria included patients at least 18 years old, diagnosis of CAD, indication for at least 1 antiplatelet drug, and willingness to participate in and sign informed consent. Exclusion criteria were a life expectancy of fewer than 6 months and current participation in another interventional clinical trial.

Treatment strategy followed contemporary guidelines, with decisions jointly made by clinicians and patients. Coronary angiography and interventional procedures were performed by experienced interventional cardiologists. Secondary prevention medications were prescribed in accordance with guideline recommendations.

The PROMISE cohort adhered to the principles of the Declaration of Helsinki and received approval from the Ethics Committee of Fuwai Hospital. Written informed consent was obtained from all participants.

### 2.2 Data Collection

The data analysed in the current study included demographic information, clinical characteristics, laboratory test results, medication administration data, and details on coronary revascularization were extracted from electronic medical records. All collected data were checked and managed by independent statisticians, maintaining confidentiality in accordance with local laws and regulations. Investigator training and regular remote monitoring of participating were carried out to ensure data quality.

### 2.3 Definition of Variables

For the definition of glucose metabolic status, diabetes mellitus was defined as fasting plasma glucose (FPG) ≥7.0 mmol/L, 2-hour postprandial plasma glucose ≥11.1 mmol/L, glycosylated hemoglobin (HbA1c) ≥6.5%, any use of glucose-lowering medications, or self-reported diabetes history. Prediabetes was determined according to fasting glucose or HbA1c thresholds, based on either the World Health Organization (WHO) criteria (FPG 6.1−< 7 mmol/L) [19], the American Diabetes Association (ADA) definition (FPG 5.6−< 7 mmol/L; HbA1c 5.7−< 6.5%) [20], or the International Expert Committee (IEC) (HbA1c 6.0−< 6.5%) [21].

Patients who did not meet the above criteria were classified as having normal glycemia according to the respective guideline definitions.

FPG were measured using the hexokinase/glucose-6-phosphate dehydrogenase method, and HbA1c was analyzed by an automated glycosylated hemoglobin analyzer (Tosoh HLC-723G8, Tokyo, Japan). Lipid profiles were determined using an automated biochemical analyzer (Hitachi 7150, Tokyo, Japan).

### 2.4 Calculation of Post-Procedural SHR

The post-procedural SHR was calculated using the formula [FPG (mmol/L)]/ [1.59 * HbA1c (%)-2.59] [2]. Fasting plasma glucose was measured from venous samples obtained on the morning following PCI. Patients were categorized into tertiles according to their SHR values (Tertile 1, Tertile 2, and Tertile 3).

### 2.5 Follow-up and Study Endpoints

In-hospital adverse outcomes were obtained through a review of electronic medical records. Long-term clinical outcomes were collected via outpatient visits, structured telephone interviews, and text messages conducted by an independent team of clinical research coordinators at 1 and 2 years after discharge. Standardized investigator training, telephone recording, and an online follow-up system were implemented to ensure uniform data collection. Two independent cardiologists adjudicated endpoint events, and any disagreement was resolved by consensus.

The primary endpoint was major adverse cardiovascular events (MACE), defined as a composite of all-cause death, non-fatal myocardial infarction (MI), any revascularization. Secondary endpoints included cardiac death and the individual components of MACE.

### 2.6 Statistical Analysis

Continuous variables were summarized as mean ± standard deviation (SD) for normally distributed data or as median (interquartile range, IQR) for skewed data. Categorical variables were presented as frequency and percentage. Baseline differences in continuous variables were compared using analysis of variance (ANOVA) or the Kruskal–Wallis test, while categorical variables were analyzed using the chi-squared test or Fisher’s exact test, as appropriate.

Kaplan–Meier survival curves were generated across SHR tertiles and compared using the log-rank test. Cox proportional hazard regression models were used to evaluate the independent association bewteen post-procedural SHR and clinical outcomes after adjusting for potential confounders. Results were reported as hazard ratios (HRs) with corresponding 95% confidence intervals (CIs). The stratified analysis was further improved on this basis.

Restricted cubic spline (RCS) regression was conducted to assess potential nonlinear relationships between post-procedural SHR and adverse outcomes.

All statistical analyses and graphical visualizations were performed using R software version 4.3.2 (R Foundation for Statistical Computing, Vienna, Austria) and SPSS version 26.0 (IBM Corp., Armonk, NY, USA). A two-tailed P value < 0.05 was considered statistically significant.

## 3. Results

### 3.1 Patients and Baseline Characteristics

The selection process of study participants is illustrated in Figure 1. Among all patients who underwent PCI during hospitalization, after further excluding participants without glycated hemoglobin or post-post-procedural fasting plasma glucose values (n = 5801) and those lost to 2-years follow-up (n = 184). Ultimately, 7,423 patients were eventually included in the current analysis, the patients were further grouped according to the tertiles of post-procedural SHR.

**Figure 1.**
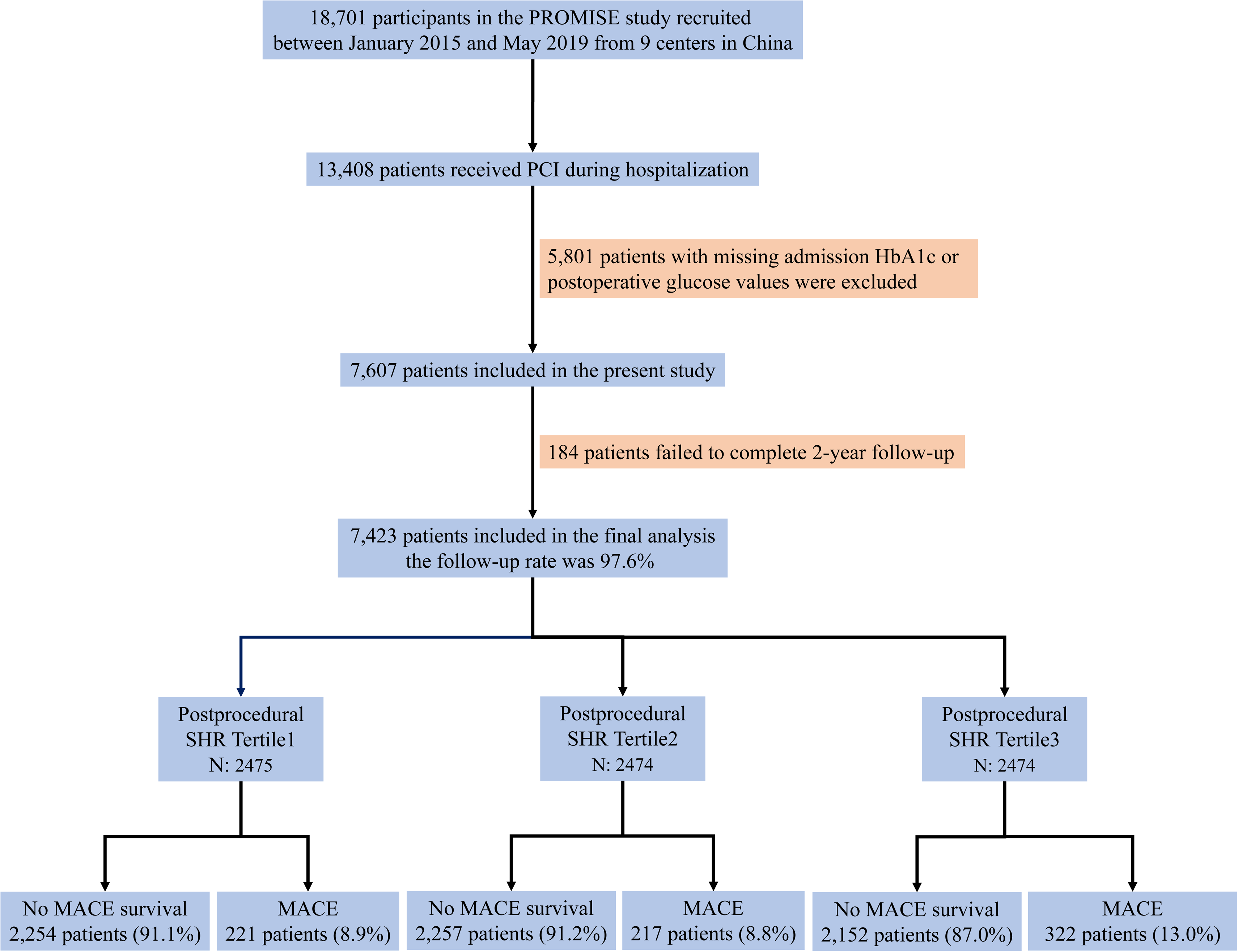
Study flowchart.

Baseline characteristics according to tertiles of post-procedural SHR are summarized in Table 1. Patients in higher SHR tertiles tended to be older and had a greater burden of comorbidities. They were more likely to present with acute myocardial infarction (AMI), to have a history of previous myocardial infarction or revascularization, and to exhibit more complex coronary disease, as indicated by higher SYNTAX scores.

### 3.2 Association between Post-Procedural SHR and Clinical Outcomes

Over a median follow-up period of 2 years, a total of 760 MACE events were recorded, including 221 (8.9%), 217 (8.8%), and 322 (13.0%) cases in the first, second, and third SHR tertiles, respectively. Kaplan–Meier survival curves demonstrated significant differences in the incidence of all adverse outcomes among SHR tertiles (log-rank P < 0.05; Figure 2).

**Figure 2.**
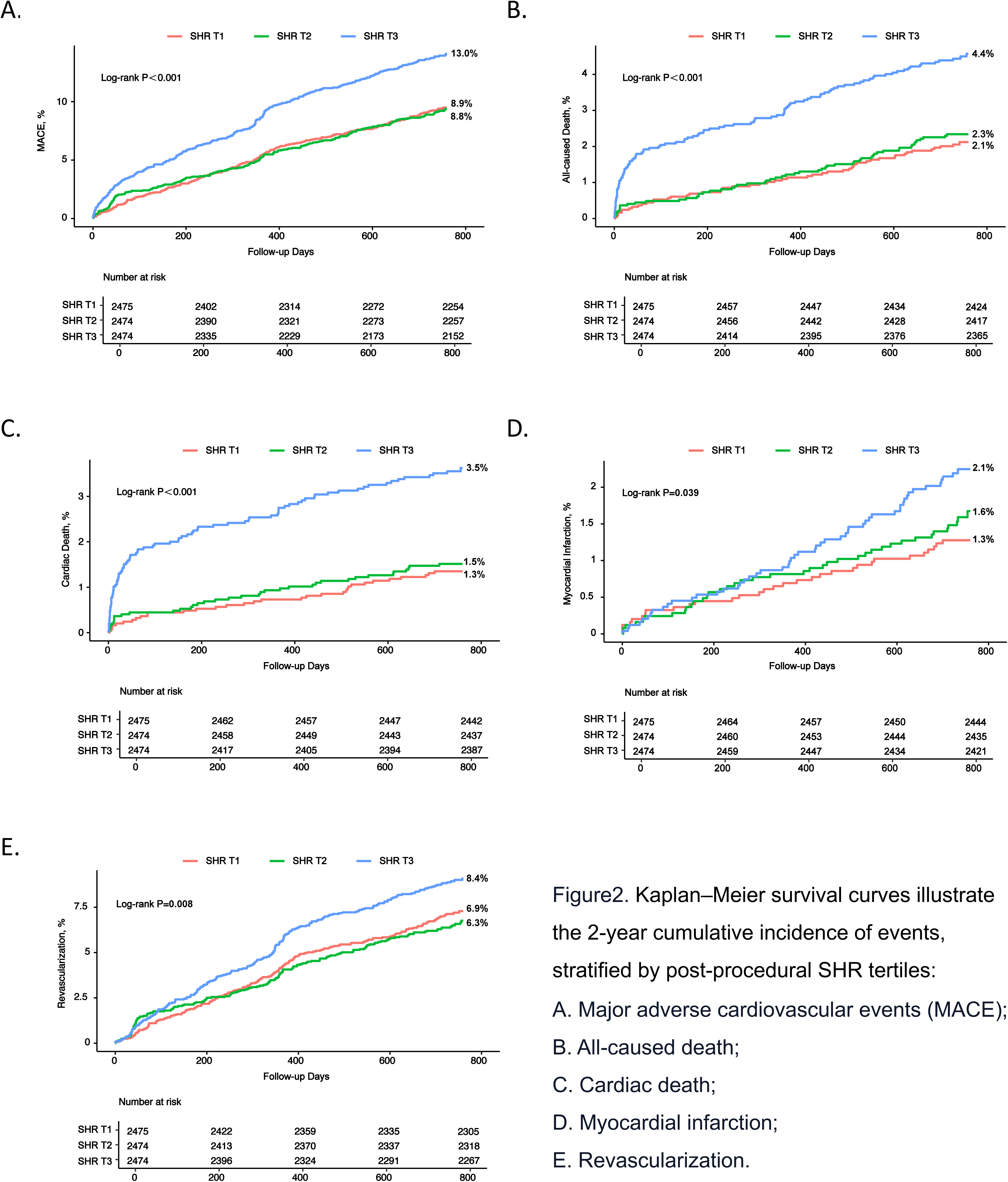
Kaplan–Meier survival curves illustrate the 2-year cumulative incidence of events, stratified by post-procedural SHR tertiles: A. Major adverse cardiovascular events (MACE); B. All-caused death; C. Cardiac death; D. Myocardial infarction; E. Revascularization.

Table2 shows the results of the Cox hazard regression, in unadjusted Cox proportional hazards analysis, patients in the highest SHR tertile had a 49% higher risk of MACE compared with the reference group. After multivariable adjustment for age, AMI, and other potential confounders, high post-procedural SHR remained an independent predictor of MACE [adjusted HR (95% CI): 1.254 (1.046–1.502), P = 0.014]. This association was mainly attributable to increased risks of all-cause death [1.531 (1.076–2.177), P = 0.018] and cardiac death [1.702 (1.111–2.607), P = 0.014], as shown in Table 3.

To further investigate potential nonlinear associations, as illustrated in Figure 3, RCS analysis revealed a J-shaped relationship between post-procedural SHR and the risk of MACE (P for overall < 0.001, P for nonlinearity = 0.102), suggesting a largely linear trend within the observed range.

**Figure 3.**
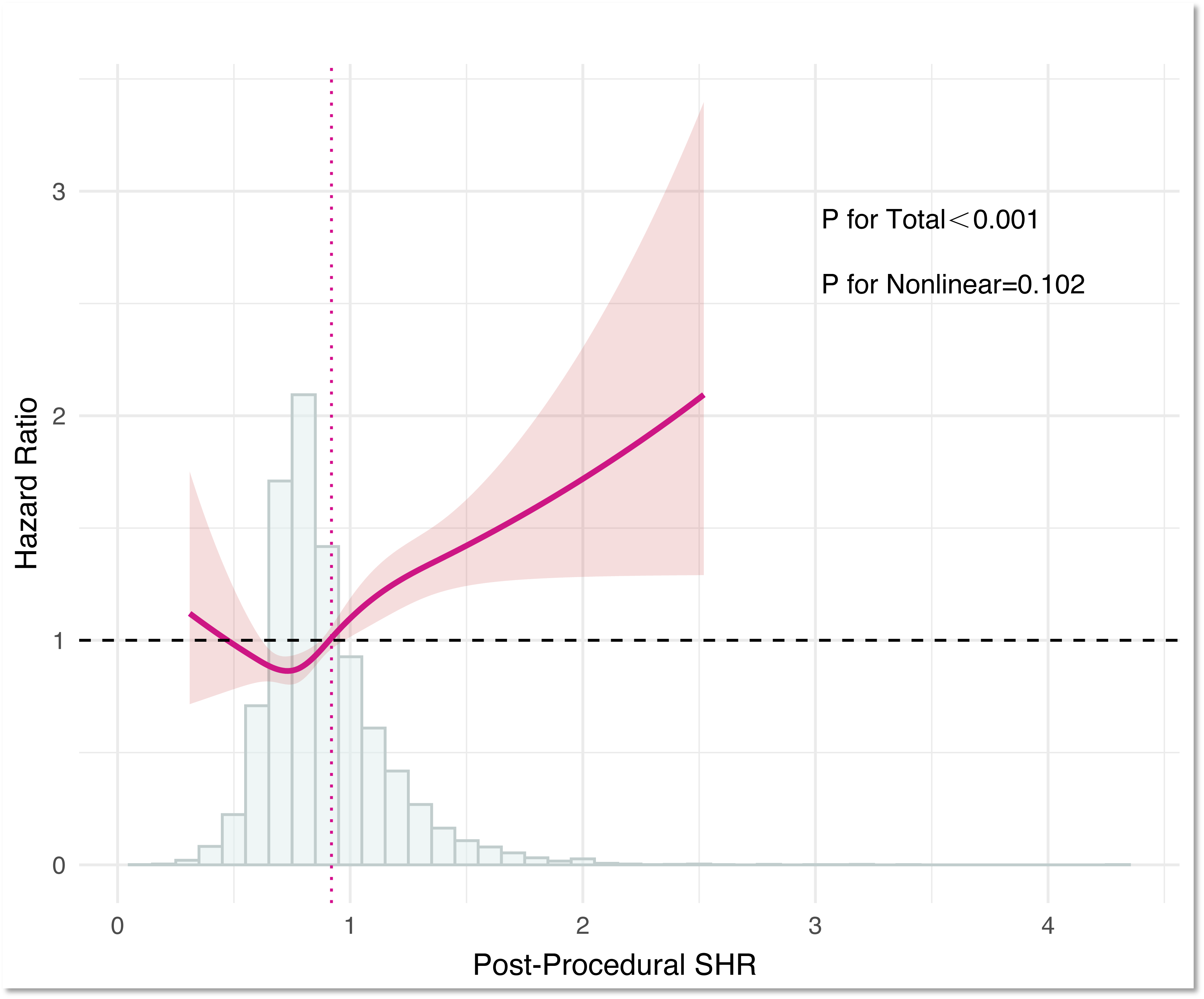
Restricted Cubic Spline with Histogram for Post-Procedural Stress Hyperglycemia Ratio.

### 3.3 Subgroup Analyses According to Glucose Metabolic Status

As shown in Table 4, the prognostic effect of post-procedural SHR was mainly observed in patients with diabetes [adjusted HR (95% CI): 1.411 (1.121–1.777), P = 0.003]. No significant association was observed among individuals with normoglycemia or prediabetes, and this result remained consistent across different international definitions of glucose metabolism status—including the IEC HbA1c-based, ADA HbA1c-based, ADA FPG-based, and WHO FPG-based criteria.

Stratified analyses suggested that glucose metabolic status might influence the association between post-procedural SHR and MACE; however, no significant interaction was observed between SHR tertiles and glucose metabolic status (all P for interaction > 0.05).

Overall, the association between SHR and MACE appeared more evident in patients with diabetes, although the modifying effect of glucose metabolic status was not statistically significant. After accounting for diabetes stage as a stratification factor in the Cox regression model, the highest SHR tertile remained an independent predictor of MACE.

## 4. Discussion

### 4.1 Major findings

In this multicenter retrospective cohort study involving 7,423 patients who underwent PCI, those in the highest tertile of post-procedural SHR experienced the greatest incidence of adverse outcomes. Elevated post-procedural SHR was independently associated with an increased risk of MACE, primarily driven by all-cause and cardiac mortality. This prognostic association was most evident among patients with diabetes, whereas no significant relationship was observed in individuals with prediabetes or normoglycemia, irrespective of the diagnostic criteria applied.

### 4.2 Differences from Previous Studies

Admission glucose levels may be markedly influenced by patients’ baseline glycemic status and thus fail to accurately reflect true stress-related hyperglycemia. The stress hyperglycemia ratio (SHR), a recently recognized metric, adjusts acute glucose levels for chronic glycemic exposure by incorporating HbA1c, thereby providing a simple and effective estimate of stress-induced hyperglycemia [2].

Previous studies have consistently demonstrated the prognostic significance of SHR across different subsets of coronary artery disease. In particular, a recent meta-analysis of 26 cohort studies reported that elevated SHR levels were significantly associated with a higher risk of major adverse cardiac and cerebrovascular events (MACCE) among patients with AMI [22]. Similar findings have also been observed in those with acute coronary syndrome (ACS) involving three-vessel disease, underscoring the robust prognostic value of SHR in acute ischemic settings [23]. However, these investigations primarily focused on stress hyperglycemia caused by the disease itself. To date, no study has evaluated the impact of post-procedural stress hyperglycemia—as reflected by SHR measured after PCI—on long-term outcomes among the broader CAD population.

Notably, Pei et al. reported that a higher postoperative SHR level was associated with increased risks of in-hospital, 90-day, and 360-day all-cause mortality after coronary artery bypass grafting (CABG) [24]. Similarly, You et al. demonstrated that greater perioperative glycemic variability predicted adverse in-hospital outcomes following CABG [25]. Extending these findings, our study is the first to show that elevated post-procedural SHR also confers significant long-term prognostic value in patients undergoing PCI—particularly among those with diabetes.

### 4.3 Mechanisms Linking Post-Procedural SHR to Adverse Cardiovascular Outcomes

Stress-induced hyperglycemia triggers excessive activation of the sympathetic nervous system and hypothalamic–pituitary–adrenal axis, resulting in elevated levels of catecholamines, cortisol, glucagon, and growth hormone. These hormones stimulate hepatic gluconeogenesis, and reduce peripheral glucose utilization, thereby promoting insulin resistance and metabolic imbalance [26]. In parallel, acute hyperglycemia enhances oxidative stress and inflammatory activation, leading to excessive generation of reactive oxygen species (ROS) and endothelial injury—key processes that accelerate atherosclerosis progression [27, 28]. In diabetic patients, stress hyperglycemia disrupts fibrinolysis by up-regulating plasminogen activator inhibitor-1 (PAI-1) and impairs antiplatelet response leading to hypercoagulability and thrombosis [29, 30].

PCI itself is a significant physiological stressor: (1) During the procedure, unavoidable endothelial injury and plaque fragmentation may trigger local or systemic inflammatory responses, characterized by the release of inflammatory mediators, recruitment of leukocytes, and activation of matrix metalloproteinases (MMPs) [15, 31]; (2) Following vessel revascularization, reperfusion injury may occur due to calcium overload, mitochondrial dysfunction, and oxidative stress, which further leads to myocardial damage [16, 32]; (3) Additionally, procedural manipulation, plaque rupture, and stent implantation can enhance platelet reactivity and thrombotic potential [17, 18].

Therefore, elevated post-procedural SHR levels may exacerbate endothelial dysfunction, oxidative stress, and a prothrombotic state, ultimately increasing the risk of subsequent adverse cardiovascular events.

### 4.4 Impact of Diabetes on Post-Procedural SHR and Cardiovascular Risk

Stratified analyses suggested that patients with diabetes may represent a high-risk population for the adverse effects of post-procedural SHR. However, no significant interaction was observed between SHR and glucose metabolic status on MACE, which may be attributable to limited sample size or the need for longer follow-up.

These findings emphasize the importance of monitoring and managing post-procedural SHR in diabetic patients, while also recognizing its contribution to adverse outcomes in the whole study population.

This finding could be explained by the presence of insulin resistance and metabolic dysregulation in patients with diabetes, which is often accompanied by chronic low-grade inflammation [33, 34]. The combined effect of these risk factors may lead to more pronounced endothelial injury and thrombotic responses following PCI. In contrast, for non-diabetic and pre-diabetic patients, post-procedural stress-induced hyperglycemia may not cause the same degree of metabolic imbalance and inflammatory response. This could explain why the increased cardiovascular risk associated with elevated SHR is most evident in diabetic patients.

## 5. Limitations

(1) Post-procedural SHR was assessed only using fasting blood glucose on the first day after the procedure, which could not dynamically capture periprocedural glucose fluctuations. (2) Information regarding periprocedural glycemic management, nutritional status, and infection was not collected, which may have influenced blood glucose levels. (3) All study participants were Chinese patients, the generalizability of these findings to other populations requires further validation.

## 6. Conclusion

Elevated post-procedural SHR is independently associated with an increased risk of adverse cardiovascular outcomes in CAD patients, particularly those with diabetes. Incorporating SHR monitoring into clinical practice could help identify high-risk individuals, enabling more tailored post-procedural management to improve patient outcomes.

## 7. Author contributions

Ying Song, Jinqing Yuan, Yaling Han, Xueyan Zhao, Jue Chen, Runlin Gao, Lei Song Participated in the study design. Weiting Cai, Lin Jiang, Jingjing Xu, Yi Yao, Na Xu, Xiaozeng Wang, Zhenyu Liu, Zheng Zhang, Yongzhen Zhang, Xiaogang Guo, Zhifang Wang, Yingqing Feng, Qingsheng Wang, Jianxin Li Participated in data analysis. Queyun Sun and Cheng Cui Wrote the manuscript. All authors participated in data interpretation and critical revision of the manuscript.

## 8. Ethics approval and consent to participate

The Institutional Review Board of Fuwai Hospital approved the study protocol, and the IRB number of this study is IRB2012-BG-006. The entire research was conducted in accordance with relevant regulations, and informed consent was obtained from all participants. The research was performed in accordance with the Declaration of Helsinki.

## 9. Consent for publication

Written informed consent for publication was obtained from all participants.

## 10. Data availability statement

The datasets generated or analyzed during the current study are not publicly available due to the participants did not agree for their data to be shared publicly but are available from the corresponding author on reasonable request.

## 11. Declaration of interests

The authors declare that they have no known competing financial interests or personal relationships that could have appeared to influence the work reported in this paper.

## 12. Acknowledgements

We thank all staff members for data collection, data entry, and monitoring as part of this study.

## 13. Funding

This work was supported by the CAMS Innovation Fund for Medical Sciences (2023-I2M-C&T-B-061) ; the National High Level Hospital Clinical Research Funding (2024-GSP-TJ-8); the National Clinical Research Center for Cardiovascular Diseases, Fuwai Hospital, Chinese Academy of Medical Sciences (NCRC2022003).

## Notes

### Competing Interest Statement

The authors have declared no competing interest.

### Clinical Trial

This prospective cohort study was observational in nature and did not involve any intervention; therefore, it was not registered in a clinical trial registry.

## Reference

1. Marik, P.E. and R. Bellomo, Stress hyperglycemia: an essential survival response! Crit Care Med, 2013. 41(6): p. e93–4.

2. Roberts, G.W., et al., Relative Hyperglycemia, a Marker of Critical Illness: Introducing the Stress Hyperglycemia Ratio. J Clin Endocrinol Metab, 2015. 100(12): p. 4490–7.

3. He, H.M., et al., Simultaneous assessment of stress hyperglycemia ratio and glycemic variability to predict mortality in patients with coronary artery disease: a retrospective cohort study from the MIMIC-IV database. Cardiovasc Diabetol, 2024. 23(1): p. 61.

4. Wang, F., et al., Combined assessment of stress hyperglycemia ratio and glycemic variability to predict all-cause mortality in critically ill patients with atherosclerotic cardiovascular diseases across different glucose metabolic states: an observational cohort study with machine learning. Cardiovasc Diabetol, 2025. 24(1): p. 199.

5. Qiao, Z., et al., High stress hyperglycemia ratio predicts adverse clinical outcome in patients with coronary three-vessel disease: a large-scale cohort study. Cardiovasc Diabetol, 2024. 23(1): p. 190.

6. Xu, W., et al., Association of stress hyperglycemia ratio and in-hospital mortality in patients with coronary artery disease: insights from a large cohort study. Cardiovasc Diabetol, 2022. 21(1): p. 217.

7. Li, X.H., et al., Predicting 28-day all-cause mortality in patients admitted to intensive care units with pre-existing chronic heart failure using the stress hyperglycemia ratio: a machine learning-driven retrospective cohort analysis. Cardiovasc Diabetol, 2025. 24(1): p. 10.

8. Li, L., et al., Stress hyperglycemia ratio and the clinical outcome of patients with heart failure: a meta-analysis. Front Endocrinol (Lausanne), 2024. 15: p. 1404028.

9. Cao, B., et al., The association between stress-induced hyperglycemia ratio and cardiovascular events as well as all-cause mortality in patients with chronic kidney disease and diabetic nephropathy. Cardiovasc Diabetol, 2025. 24(1): p. 55.

10. Chen, Y., et al., Assessment of stress hyperglycemia ratio to predict all-cause mortality in patients with critical cerebrovascular disease: a retrospective cohort study from the MIMIC-IV database. Cardiovasc Diabetol, 2025. 24(1): p. 58.

11. Zhang, Y., et al., Association between the stress hyperglycemia ratio and mortality in patients with acute ischemic stroke. Sci Rep, 2024. 14(1): p. 20962.

12. Yan, F., et al., Association between the stress hyperglycemia ratio and 28-day all-cause mortality in critically ill patients with sepsis: a retrospective cohort study and predictive model establishment based on machine learning. Cardiovasc Diabetol, 2024. 23(1): p. 163.

13. Cheng, S., et al., Association between stress hyperglycemia ratio index and all-cause mortality in critically ill patients with atrial fibrillation: a retrospective study using the MIMIC-IV database. Cardiovasc Diabetol, 2024. 23(1): p. 363.

14. Li, L., et al., Prognostic significance of the stress hyperglycemia ratio in critically ill patients. Cardiovasc Diabetol, 2023. 22(1): p. 275.

15. Inoue, T., et al., Vascular inflammation and repair: implications for re-endothelialization, restenosis, and stent thrombosis. JACC Cardiovasc Interv, 2011. 4(10): p. 1057–66.

16. Fröhlich, G.M., et al., Myocardial reperfusion injury: looking beyond primary PCI. Eur Heart J, 2013. 34(23): p. 1714–22.

17. Alexopoulos, D., et al., Peri-Procedural Platelet Reactivity in Percutaneous Coronary Intervention. Thromb Haemost, 2018. 118(7): p. 1131–1140.

18. Gawaz, M., et al., Platelet activation and coronary stent implantation. Effect of antithrombotic therapy. Circulation, 1996. 94(3): p. 279–85.

19. Colagiuri, S., Definition and Classification of Diabetes and Prediabetes and Emerging Data on Phenotypes. Endocrinol Metab Clin North Am, 2021. 50(3): p. 319–336.

20. Report of the expert committee on the diagnosis and classification of diabetes mellitus. Diabetes Care, 2003. 26 Suppl 1: p. S5-20.

21. Gillett, M.J., International Expert Committee report on the role of the A1c assay in the diagnosis of diabetes: Diabetes Care 2009; 32(7): 1327-1334. Clin Biochem Rev, 2009. 30(4): p. 197–200.

22. Karakasis, P., et al., Prognostic value of stress hyperglycemia ratio in patients with acute myocardial infarction: A systematic review with Bayesian and frequentist meta-analysis. Trends Cardiovasc Med, 2024. 34(7): p. 453–465.

23. Zhang, Y., et al., Effects of the stress hyperglycemia ratio on long-term mortality in patients with triple-vessel disease and acute coronary syndrome. Cardiovasc Diabetol, 2024. 23(1): p. 143.

24. Pei, Y., et al., Stress hyperglycemia ratio and machine learning model for prediction of all-cause mortality in patients undergoing cardiac surgery. Cardiovasc Diabetol, 2025. 24(1): p. 77.

25. You, H., et al., Effect of glycemic control and glucose fluctuation on in-hospital adverse outcomes after on-pump coronary artery bypass grafting in patients with diabetes: a retrospective study. Diabetol Metab Syndr, 2023. 15(1): p. 20.

26. Shamoon, H., R. Hendler, and R.S. Sherwin, Synergistic interactions among antiinsulin hormones in the pathogenesis of stress hyperglycemia in humans. J Clin Endocrinol Metab, 1981. 52(6): p. 1235–41.

27. Monnier, L., et al., Activation of oxidative stress by acute glucose fluctuations compared with sustained chronic hyperglycemia in patients with type 2 diabetes. Jama, 2006. 295(14): p. 1681–7.

28. Yamamoto, Y. and H. Yamamoto, *RAGE-Mediated Inflammation,* Type 2 Diabetes, and Diabetic Vascular Complication. Front Endocrinol (Lausanne), 2013. 4: p. 105.

29. Worthley, M.I., et al., The deleterious effects of hyperglycemia on platelet function in diabetic patients with acute coronary syndromes mediation by superoxide production, resolution with intensive insulin administration. J Am Coll Cardiol, 2007. 49(3): p. 304–10.

30. Violi, F., et al., Nutrition, Thrombosis, and Cardiovascular Disease. Circ Res, 2020. 126(10): p. 1415–1442.

31. Tucker, B., et al., Inflammation during Percutaneous Coronary Intervention-Prognostic Value, Mechanisms and Therapeutic Targets. Cells, 2021. 10(6).

32. Hausenloy, D.J. and D.M. Yellon, Myocardial ischemia-reperfusion injury: a neglected therapeutic target. J Clin Invest, 2013. 123(1): p. 92–100.

33. Rohm, T.V., et al., Inflammation in obesity, diabetes, and related disorders. Immunity, 2022. 55(1): p. 31–55.

34. Accili, D., Z. Deng, and Q. Liu, Insulin resistance in type 2 diabetes mellitus. Nat Rev Endocrinol, 2025. 21(7): p. 413–426.

